# Emergence and Spread of Artemisinin-Resistant Malaria in Zambia

**DOI:** 10.64898/2026.06.04.26354343

**Authors:** Mulenga Mwenda, Rafael Oliveira, Brenda Mambwe, Carol Chiyesu, Bernd Bohmeier, Karolina Mosler, Miller Phiri, Andy Sinyoolo, Virginia Chiposa, Tupilwe Namonje, Memory Munsanje, Matthews Ilunga, Chitundu Chirwa, Innocent Mwape, Donald Mumba, Romain Coppée, Maria-Alexandra Stoica, Maria Isabel Veiga, Chris Drakeley, Richard Pearson, Robert Verity, Jacob Chirwa, Frank P. Mockenhaupt, Welmoed Van Loon, Silvia Portugal, Edgar Simulundu, Stephen Bwalya, John M. Miller, Roma Chilengi, Charles Fanaka, Daniel J. Bridges, Moonga Hawela, Jason A. Hendry

## Abstract

**Background:** Artemisinin derivatives are central to first-line treatment of both uncomplicated and severe *Plasmodium falciparum* malaria. Emerging artemisinin partial resistance in East Africa threatens to spread across the continent.

**Methods:** In two cross-sectional studies in Zambia in 2024, we genotyped the artemisinin resistance-associated gene *Pfkelch13*. In Kaoma, western Zambia, we evaluated the percentage of patients with day-3 parasite positivity following treatment with artemisinin-based combination therapy, and *ex vivo* parasite susceptibility to dihydroartemisinin (the active metabolite of artemisinin). We also assessed longitudinal changes in *Pfkelch13* mutation prevalence in Kaoma using isolates collected from 2018 through 2026.

**Results:** We identified a novel mutation, *Pfkelch13* A724E, in 52% (113 of 217) of isolates from Western Province, 51% (94 of 184) of isolates from North-Western Province, and 11.7% (229 of 1,949) of isolates country-wide. In Kaoma, 28% (21 of 75) of patients carrying *Pfkelch13* A724E mutant parasites before treatment were parasite positive on day 3, compared with 0% (0 of 23) of patients with the wild-type allele (P=0.003). Within day-3 positive patients, the proportion of A724E mutant parasites increased significantly after treatment (P = 0.013). The prevalence of *Pfkelch13* A724E in Kaoma increased steadily from 0% (95% confidence interval [CI], 0 to 22%) in 2018 to 79% (95% CI, 73 to 85%) in 2026.

**Conclusions:** A novel *Pfkelch13* mutation conferring partial resistance to artemisinin is spreading in Zambia. Additional clinical evaluations are urgently needed in the region. (Funded by the Gates Foundation, INV-048316).

## Introduction

The plant-derived molecule artemisinin, with its unique endoperoxide bridge, is central to modern treatment of *Plasmodium* (*P*.) *falciparum* malaria^1^. Artemisinin-based combination therapies coformulate a rapidly acting derivative of artemisinin (artemether, dihydroartemisinin, or artesunate) with a longer-acting partner drug (such as lumefantrine, amodiaquine, or piperaquine), and are the recommended first-line treatment for uncomplicated *P. falciparum* cases; while parenteral artesunate is recommended for severe cases^2^.

These treatments, however, are endangered by the spread of artemisinin partial resistance. Artemisinin partial resistance is defined clinically as delayed parasite clearance following treatment; or *in vitro* as increased parasite survival following artemisinin derivative exposure^3^. Molecular and association studies identified non-synonymous point mutations in the beta-propeller-encoding domain of *Pfkelch13* as the primary cause of this resistance^4,5^. First detected near the Thailand-Cambodia border in 2008^6,7^, parasites with artemisinin partial resistance subsequently acquired resistance to partner drugs, and spread throughout the Greater Mekong Subregion^8,9^. A decline in the clinical efficacy of specific artemisinin-based combination therapies followed^10^, necessitating revisions to national treatment policies.

Now in East Africa, strong evidence that artemisinin partial resistance has emerged is accumulating. In Rwanda, the marker *Pfkelch13* R561H arose in 2014, and was shown to confer clinical and *in vitro* artemisinin partial resistance^11,12^. Since then, four World Health Organization (WHO)-validated markers of artemisinin partial resistance^13^ have been identified in the region: *Pfkelch13* R561H in Rwanda^11,12^, Uganda^14^ and Tanzania^15^; C469Y and A675V, both only in Uganda^14,16^; and R622I in Eritrea^17^, Ethiopia^18^ and Sudan^19^. Genetic investigations revealed that these mutations emerged locally, and all but R622I show evidence of clonal spread. Though some have increased to moderate prevalence in their respective geographies^20^, the clinical efficacy of artemisinin-based combination therapies remain largely high, likely due to the absence of significant partner drug resistance.

In the Southern African country of Zambia, previously reported resistance-associated mutations in *Pfkelch13* have been detected, however at low prevalence, and without local evidence of resistance^21,22^. Here, we conducted a cross-sectional genotyping study that resulted in the identification of a hitherto unreported *Pfkelch13* mutation, A724E, at high prevalence over a vast area of western Zambia. This prompted a clinical study which found that A724E is associated with artemisinin partial resistance.

## Methods

### Study design and participants

To investigate the spatial distribution of artemisinin partial resistance markers, we analysed samples from two previously unpublished cross-sectional studies conducted in Zambia between April and October 2024: a community-based national malaria indicator survey and a health facility-based molecular marker study. The national malaria indicator survey was a two-stage cluster-randomised population study designed to generate nationally-representative estimates of *P. falciparum* epidemiological metrics. In randomly selected households, children aged 6 to 72 months were recruited. The molecular study was a multi-site observational study conducted at 100 health facilities across all 10 provinces of Zambia designed to estimate molecular marker prevalence. Patients aged 6 months or older presenting with malaria symptoms were recruited. In both studies, diagnosis was by rapid diagnostic test and participants were enrolled if their informed written consent was obtained or, if a child, their assent was obtained along with consent from the parent or guardian.

To characterise clinical and *ex vivo* antimalarial susceptibility phenotypes, we conducted a single-site observational study at the Kaoma District Hospital in Western Province, Zambia, in April 2026. Malaria is seasonal in Kaoma, with an annual incidence of 943 cases per 1,000 population in 2025. Patients aged 12 months or older presenting for routine malaria diagnosis were recruited if they had a positive rapid diagnostic test and microscopically confirmed *P. falciparum* infection with ≥10,000 parasites per cubic millimeter. Patients with self-reported prior treatment for malaria were excluded. Patients were prescribed an unsupervised 3-day course of artemether-lumefantrine according to national treatment guidelines. Patients were enrolled if their informed written consent and willingness for a follow-up visit on day 3 post-treatment was obtained or, if a child, their assent was obtained along with consent from a parent or guardian.

The studies were approved by the University of Zambia Biomedical Research Ethics Committee (5055-20241, 7840-2026) and Tropical Diseases Research Centre Ethics Review Committee (TDREC/036/04/2023).

### Sample collection

In the cross-sectional studies, dried-blood spot filter papers were collected by finger prick for molecular analysis.

For the clinical study at Kaoma District Hospital, venous blood samples (<5mL) were collected on day 0 for *ex vivo* analysis of parasite drug susceptibility. For patients returning on day 3, parasite positivity was assessed by microscopic examination of thick and thin blood smears by two independent microscopists. Dried-blood spots were collected on day 0 and day 3 for molecular analysis.

### Molecular analysis

DNA was extracted from dried blood spots using the QIAamp DNA MiniKit (Qiagen, 51306). *P. falciparum* positivity was evaluated using conventional^23^ or quantitative polymerase-chain-reaction (PCR)^24^. Targeted nanopore sequencing of *P. falciparum* was performed at the National Malaria Elimination Center in Lusaka, Zambia using the previously published NOMADS-MVP protocol^25^. This protocol includes an amplicon covering *PfKelch13* from codon 383 to the carboxyl-terminus (codon 727). Sequencing data was processed using MinKNOW and the Nomadic pipeline (v0.7.0)^26^. Samples with ≥100-fold mean coverage over *Pfkelch13*, no evidence of sequencing contamination, and no evidence of sample duplication were included in downstream analysis.

We genotyped seven microsatellite loci flanking *Pfkelch13* from -31.9kbp to 72.3kbp using previously described primers^14,27^. Only samples with all seven loci successfully genotyped were included in downstream analyses. Details are available in the Supplementary Appendix (Table S1).

### *Ex vivo* drug susceptibility assays

*Ex vivo* parasite susceptibility to dihydroartemisinin and lumefantrine was evaluated using standard 72-hour half-maximal inhibitory concentration assays, as previously described^28^. Venous blood samples were collected from patients at Kaoma District Hospital and stored at 4°C overnight before assays were performed in duplicate. Growth inhibition was evaluated by fitting a four-parameter Hill equation to the dose-response of both replicates jointly, after normalisation to untreated controls.

### *In silico* structural modelling

The A724E mutation was introduced into the wildtype AlphaFold^29^ K13 structure (Q8IDQ2) using the PyMOL mutagenesis function. A 200 nanosecond molecular dynamics simulation of residues 350 to 727 was performed using GROMACS^30^, as described in the Supplementary Appendix.

### Statistical analysis

We computed the prevalence of observed *Pfkelch13* mutations treating heterozygous genotypes as mutants. Microsatellite haplotypes were hierarchically clustered based on pairwise Hamming distance. Expected heterozygosity was used to measure microsatellite genetic diversity and differences were evaluated with permutation tests. Univariable analyses used the Mann-Whitney U test, Wilcoxon signed-rank test, Chi-squared test, or Fisher’s exact test, as appropriate. Multivariable analyses used a logistic regression model with parameters estimated by Firth’s penalised maximum likelihood (to handle zero counts)^31^. Statistical analyses were performed using statsmodels (v0.14) and scipy (v1.16) libraries in Python (v3.13).

## Results

### Study samples and patients

A total of 2,614 participants were enrolled in the household survey, and 6,858 participants with malaria symptoms in the health facility-based study. Based on rapid diagnostic test and PCR positivity, we sequenced 2,399 (25%) of the 9,472 isolates collected. Of these, 449 (19%) were excluded due to low sequencing coverage, evidence of sequencing contamination, or evidence of sample duplication. For the remaining 1,950 isolates, the genotype of *Pfkelch13* was assessed (Table S2).

### Genotyping of *Pfkelch13*

Across Zambia, we observed 8 non-synonymous mutations in *Pfkelch13* at a prevalence of ≥0.5% in the 1,950 analysed isolates (Table S3, Table S4). These included the WHO validated artemisinin resistance marker *Pfkelch13* P574L in 11 isolates (0.5%; 95% CI, 0.3 to 0.9%), and the WHO candidate marker P441L in 58 isolates (2.7%; 2.1 to 3.5%). Both markers had been previously reported in Zambia in a 2023 cross-sectional study, and at a similar prevalence^21^. Notably, the highest prevalence *Pfkelch13* mutation was the non-synonymous change of alanine to glutamic acid at codon 724 (A724E). *Pfkelch13* A724E was identified in 229 (11.7%) of 1,949 isolates (95% CI, 10.4 to 13.3%).

Geographically, *Pfkelch13* A724E was at high prevalence across western Zambia, and near absent in the east (Figure 1). It was detected in 18 of 51 districts in which ten or more isolates were sequenced, and 17 (94%) of these were west of the centrally located capital city Lusaka. In Western Province, *Pfkelch13* A724E was detected in 113 (52%) of 217 isolates (95% CI, 45 to 59%); and in North-Western Province, in 94 (51%) of 184 isolates (95% CI, 44 to 59%). In other provinces, *Pfkelch13* A724E was at a much lower prevalence or absent (Table S5).

**Figure 1.**
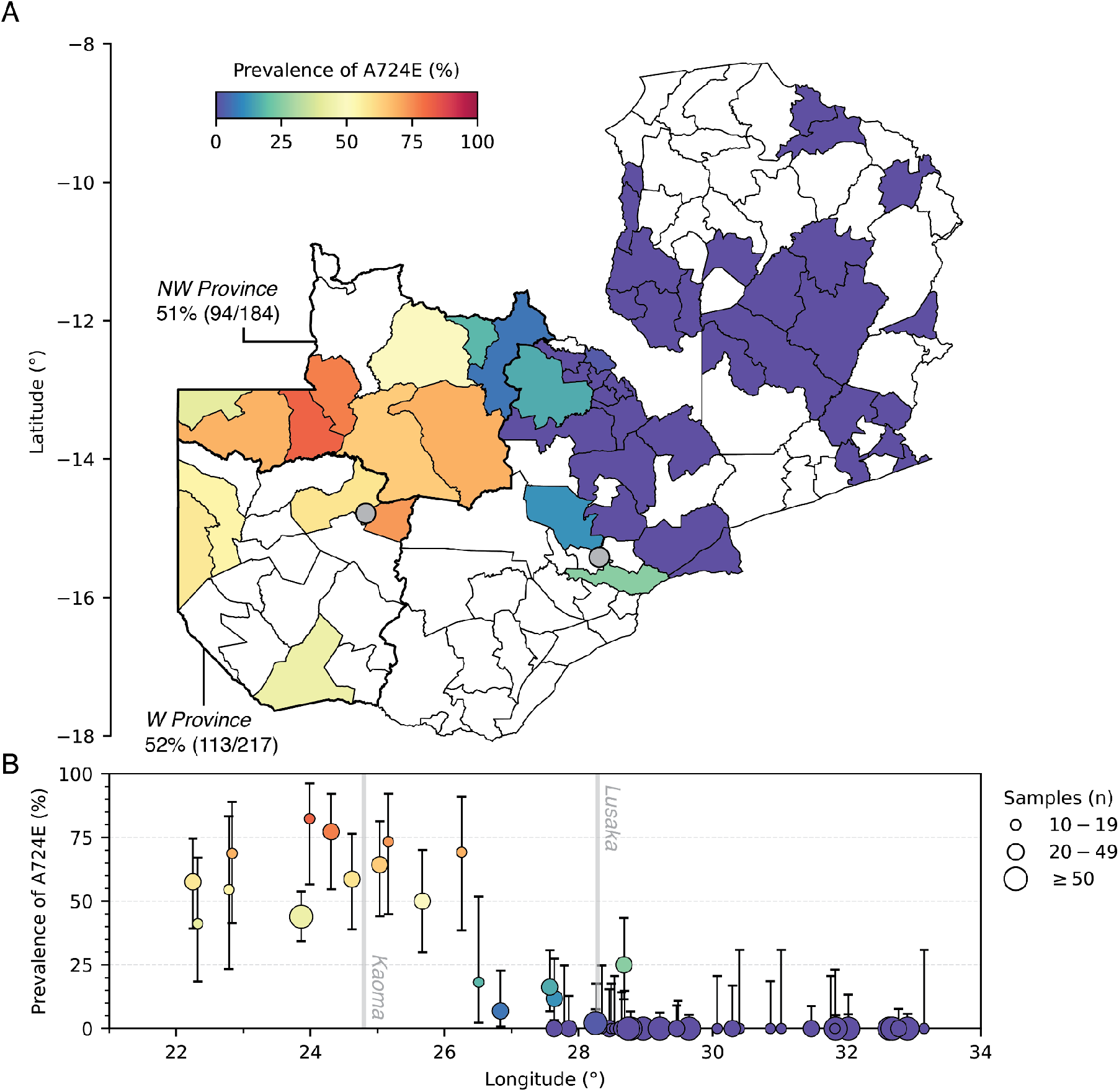
Prevalence of *Pfkelch13* A724E in 2024 across Zambia. Panel A shows the prevalence of *Pfkelch13* A724E in all districts with ten or more isolates sequenced. A total of 1,843 isolates across 51 districts are represented. Lusaka and Kaoma are marked with grey points. Panel B shows *Pfkelch13* A724E prevalence for districts in Panel A plotted against the longitude of their centroids. Whiskers are 95% CIs and point sizes represent the number of isolates sequenced.

**Figure 2.**
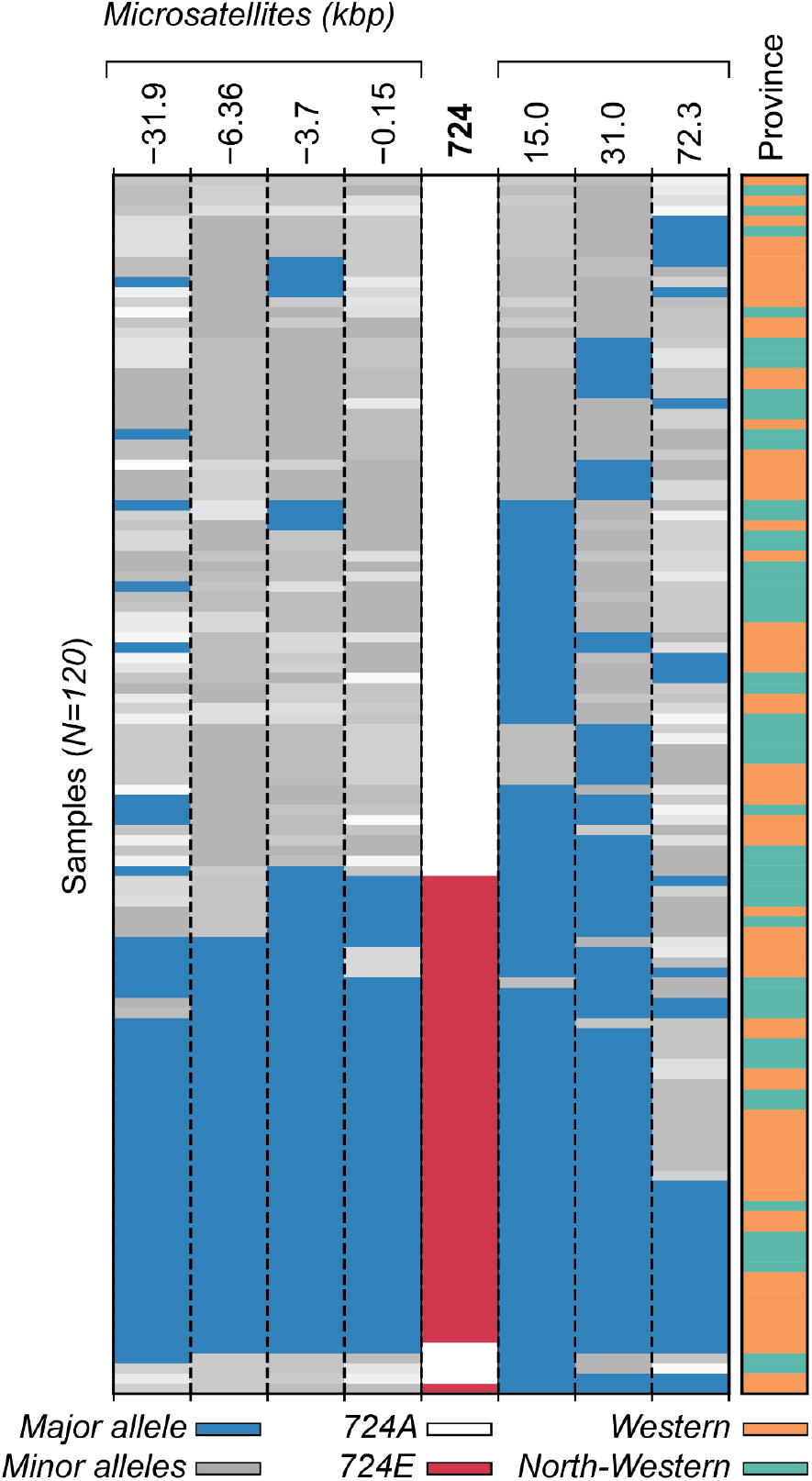
Microsatellite haplotypes flanking *Pfkelch13*. Alleles for *N=120* isolates and seven microsatellites flanking *Pfkelch13* are shown (-31.9kbp to 72.3kbp). For each microsatellite the major allele is blue and other alleles are in greyscale. *PfKelch13* A724E is in red. Rows are hierarchically clustered on haplotype hamming distance. Note a common major haplotype (-31.9kbp to 31.0kbp) in *Pfkelch13* A724E isolates across both provinces.

### Prior reporting and structural effects of *Pfkelch13* A724E

A724E changes the antepenultimate amino acid of *Pfkelch13* and, to our knowledge, has never been previously detected in Zambia or elsewhere (Supplementary Appendix). Alanine 724 is within the first propeller motif of the beta-propeller domain, in a restricted three-dimensional space (Figure S1). Structural modelling *in silico* revealed that A724E causes steric hindrance and destabilises the *Pfkelch13* protein, consistent with a partial loss of function.

### Origin of *Pfkelch13* A724E

To investigate the origin of the *Pfkelch13* A724E mutation in Zambia, we genotyped seven flanking microsatellites in 120 randomly selected clonal isolates from Western Province (n=67) and North-Western Province (n=53). The mean heterozygosity across microsatellites was significantly lower for A724E isolates than wild-type isolates (mean *H*_*e*_ 0.24 vs. 0.79, mean-difference permutation test P < 0.001); with equivalent results obtained when isolates were stratified by province (Table S6, Table S7). This was driven by a common flanking haplotype spanning from -31.9kbp to 31kbp, which accounted for 32 (68%) of 47 A724E isolates, and was present in both Western (18 of 25 A724E isolates) and North-Western Province (14 of 22 A724E isolates) (Figure 3). These data indicate that *Pfkelch13* A724E originated once in Zambia, and then spread quickly across the western provinces.

**Figure 3.**
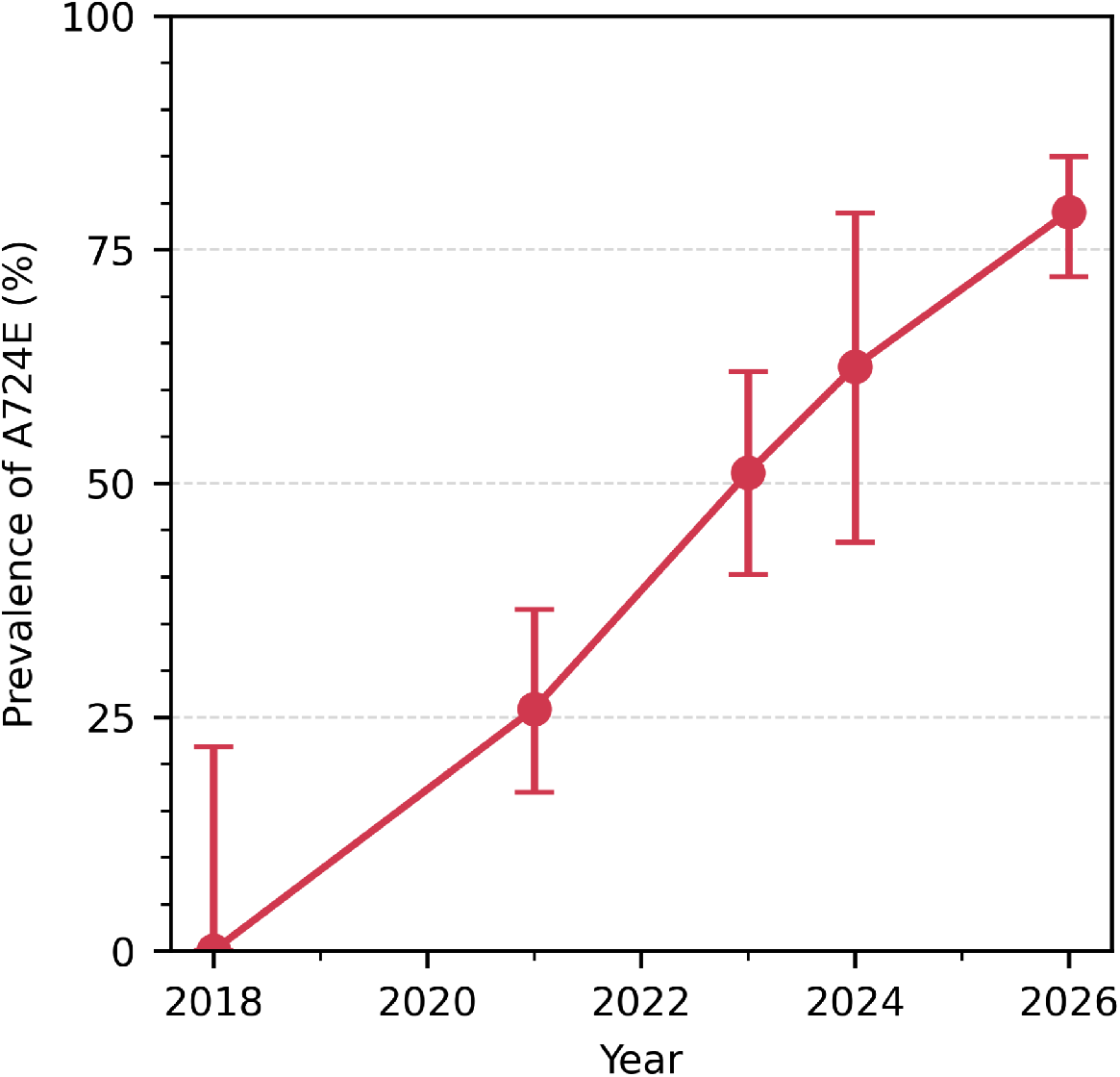
Prevalence of *Pfkelch13* A724E from 2018 through 2026 in Kaoma, Zambia. A total of *N=386* isolates are included. Data are available in Table S8. Whiskers show 95% CI.

To better understand the recent history of *Pfkelch13* A724E, we sequenced archival isolates from 2018, 2021 and 2023, which were collected as part of previous studies conducted in Kaoma, Western Province (Figure 3). From 2018 through 2026, the prevalence of A724E increased, in absolute terms, by an average of 10.2 percentage points per year. We estimated a positive selection coefficient for A724E of 0.49 (95% CI, 0.38 to 0.60; Figure S2).

### Clinical effects of *Pfkelch13* A724E

At Kaoma District Hospital, a total of 188 patients with uncomplicated *P. falciparum* infection were enrolled and prescribed the nationally-recommended, 3-day course of artemether-lumefantrine (days 0 to 2), of which 103 (54.8%) returned for a follow-up visit post-treatment (on day 3). There were no statistically significant differences observed in baseline characteristics or A724E prevalence between patients who returned versus those lost to follow-up (Table S9). Of the 103 patients who returned, 21 (20.4%) carried microscopically detectable parasites and were classified as day-3 positive (Figure 4A). In these 21 patients, post-treatment parasitemia was at least ten-fold lower than pre-treatment (Figure S3).

**Figure 4.**
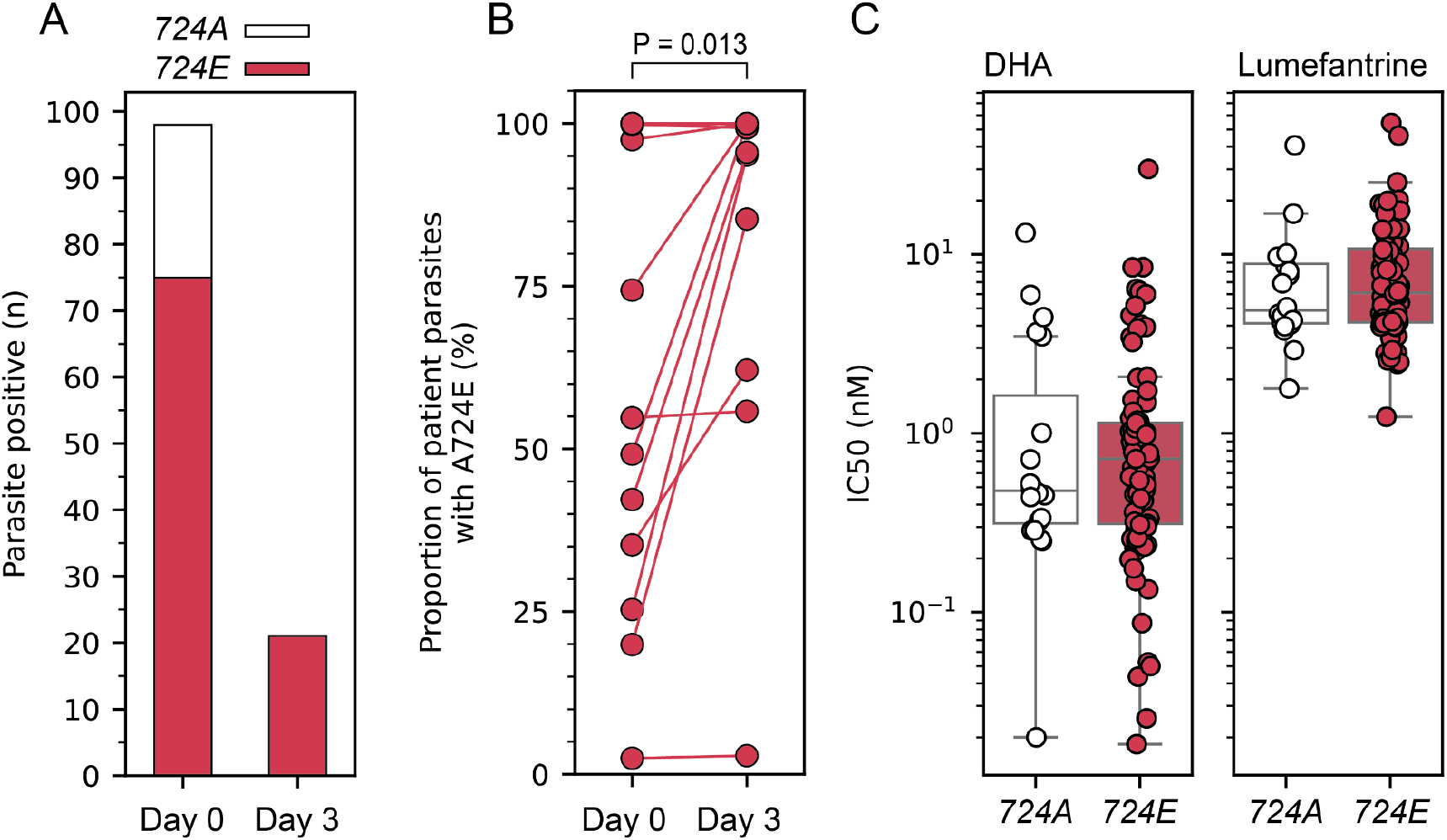
Clinical and *ex vivo* effects of *Pfkelch13* A724E. Panel A shows a cohort of *N=98* patients with *P. falciparum* infection according to their pre-treatment *Pfkelch13* genotype on day 0 and, for those that were still microscopically positive, on day 3 post-treatment. 724A is wildtype, in white; 724E is mutant, in red. Note all day-3 positive patients carried the mutant allele pre-treatment. Panel B shows the within-patient proportion of parasites with the A724E mutation pre-treatment (day 0) and post-treatment (day 3) for *N=17* day-3 positive patients with successful day 3 sequencing. Two patients with similar A724 proportions before and after treatment carried other *Pfkelch13* mutations (Table S13). Difference was significant by Wilcoxon signed-rank test. Panel C shows *ex vivo* half-maximal inhibitory concentration (IC50) distributions for dihydroartemisinin (DHA) and lumefantrine. Differences were not significant by Mann-Whitney U test.

For returning patients, 98 (95%) of pre-treatment isolates were successfully genotyped. In a multivariable analysis (STable 10), day-3 positivity was significantly associated with pre-treatment parasite density (odds ratio 5.7; 95% CI, 1.4 to 30.5) and the presence of A724E (odds ratio 13.7; 95% CI, 1.6 to 1803) (Table 1, Table S11). Both associations remained when 16 patients who carried other *Pfkelch13* mutations were excluded (STable 12, STable 13). All 21 day-3 positive patients carried A724E before treatment; corresponding to a 28% (21 of 75) day-3 positivity rate for A724E carriers. All 23 patients carrying the wildtype allele before treatment were negative on day 3.

**Table 1.** Associations between covariates and day-3 positivity among 98 patients with uncomplicated *P. falciparum* malaria who presented at Kaoma District Hospital, Western Province, in April 2026. Allowing for age, sex, fever, and initial parasite density, patients carrying the *Pfkelch13* A724E mutation are 13.7 (95% CI, 1.6 – 1802.7) fold more likely to be day-3 positive. Initial parasite density was also significantly associated with day-3 positivity. Estimates of coefficients and odds ratios are from multivariable logistic regression fit by maximising the likelihood with Firth’s penalty^31^. P values indicate significance of association by a Wald test.

| Covariate | Coefficient (SE) | Odds Ratio (95% CI) | P |
| --- | --- | --- | --- |
| Age | -0.01 (0.03) | 1.0 (0.9 – 1.0) | 0.73 |
| Sex | -0.8 (0.54) | 0.5 (0.1 – 1.3) | 0.146 |
| Fever | 0.28 (0.6) | 1.3 (0.4 – 4.3) | 0.655 |
| Parasite density [log10] | 1.75 (0.75) | 5.7 (1.4 – 30.5) | 0.016 |
| *Pfkelch13* A724E | 2.61 (1.39) | 13.7 (1.6 – 1802.7) | 0.011 |

*Pfkelch13* was successfully genotyped post-treatment for 17 (81%) of 21 patients with persisting parasitemia. For these, we could analyse the proportion of total parasites within each patient that were A724E mutants before and after treatment (Figure 4B). We observed a statistically significant increase in the proportion of A724E mutant parasites within patients after treatment (P = 0.013, Wilcoxon signed-rank test). Before treatment 9 of 17 patients carried a mixture of A724E mutant and wildtype parasites, with a median A724E proportion of 42% (IQR 25% to 55%); after treatment the median A724E proportion in these patients was 95% (IQR 62% to 100%). Only two patients had similar A724E proportions before and after treatment, and these patients also carried other *Pfkelch13* mutations (Table S14).

### *Ex vivo* effects of *Pfkelch13* A724E

We generated *ex vivo* antimalarial drug susceptibility data for 108 (57%) of 188 isolates from Kaoma (Figure 4C). We observed no significant differences between wildtype and A724E isolates in half-maximal inhibitory concentrations for dihydroartemisinin or lumefantrine (Table S15).

## Discussion

We have identified a previously unreported mutation in the artemisinin-resistance associated gene *Pfkelch13*, namely A724E, circulating at high prevalence across western Zambia. The mutation has clinical relevance; it is associated with increased day-3 parasite positivity and is selected for within-patients following artemether-lumefantrine treatment. Concerningly, genetic analyses indicated that A724E originated once, increased rapidly in prevalence, and by 2024 already spanned a vast geographical area. Combined, these findings are consistent with a novel artemisinin resistance marker undergoing a hard selective sweep in Africa.

The location and extent of A724E are notable. In our study, it exceeded 50% prevalence in Zambia’s Western and North-Western Province, which have a combined area of 252 thousand kilometers squared – larger than Uganda, or nearly ten-times the size of Rwanda. Other highly prevalent *Pfkelch13* mutations have been observed in Africa, but rarely, and over much smaller areas^20^. Moreover, prevalent *Pfkelch13* mutations in Africa have, to date, been largely confined to countries in East Africa. Our results show this is no longer the case – artemisinin partial resistance has emerged in Southern Africa.

Furthermore, A724E appears to have increased in prevalence more rapidly than previous *Pfkelch13* mutations reported in Africa. From 2018 through 2026, it increased in absolute terms by 10.2% per year, compared to 3.4% for R561H in northern Rwanda, or 2.8% for C469Y in northern Uganda^20^. Moreover, while these other *Pfkelch13* mutations appear to be equilibrating at intermediate prevalence, A724E appears to be heading towards fixation.

Among existing artemisinin resistance markers, A724E is located uniquely close to the carboxyl-terminus of *Pfkelch13*. Widely used genetic surveillance methods for *Pfkelch13* often focus on the core of the beta-propeller-encoding domain, and do not include codon 724^4,32^. This explains, in part, the lack of earlier detection^21^. Whole-genome sequencing would enable detection of A724E, but for routine surveillance is cost-prohibitive. We detected A724E using targeted nanopore sequencing^25^.

Our study has limitations. Although our cross-sectional study was country-wide, both our longitudinal and clinical results derive from a single district in Western Province, and the degree to which they generalise across the wider region is unknown. Given that our microsatellite analysis indicated a single origin for A724E, some commonality is expected. We also highlight the high rate of loss to follow-up and lack of supervised treatment in assessing day-3 positivity. Although no bias was observed in patient baseline characteristics, differences could exist in unmeasured variables. Non-adherence could inflate the rate of day-3 positivity, however our comparison between *Pfkelch13* genotypes should be robust to this possibility. While we did not observe a significant effect of A724E in *ex vivo* growth inhibition assays, this is not unusual, as these assays are notoriously insensitive^4,28,33^.

Action should be taken to ensure the impact of A724E on malaria treatment, control, and transmission is minimised. Further evaluations of the clinical consequences of A724E are needed. In parallel, clinical findings should be corroborated through gene editing and culture adaptation of field parasites. To guide a regional response, the full geographical extent of A724E should be determined through genotyping studies in neighbouring countries with suitable assays.

Potential public health responses to artemisinin-resistance in Africa have been outlined by the WHO^3^. The primary partner drug in the region is lumefantrine, which will now bear a greater clinical burden, and a marker of lumefantrine resistance has been recently identified in Uganda^34^. Multiple first-line therapies may dampen any partner drug resistance, should it arise in Southern Africa, and many tools still exist to which the parasites will remain fully susceptible, such as those targeting the vector and vaccines. Recently, novel non-artemisinin antimalarials have had promising Phase III trial results^35^. They may soon be needed.

## Data Availability

All data produced in the present study are available upon reasonable request to the authors.

## Acknowledgements

We thank all patients, guardians, health workers, and operational staff who participated in and supported these studies; the National Malaria Elimination Centre and Zambia National Public Health Institute for facilitating our work and providing laboratory space; and Estée Török for support and scientific feedback throughout. This work was funded by the Gates Foundation (INV-048316).

