## Supplementary Appendix for "Emergence and Spread of Artemisinin-Resistant Malaria in Zambia"

### Emergence and Spread of Artemisinin-Resistant Malaria in Zambia
*Supplementary Appendix*

**Author Contributions** 3
**Supplementary Methods** 3
Nanopore sequencing 3
Microsatellite genotyping 3
Molecular dynamics simulations 4
Archival sample sequencing 5
**Supplementary Information** 6
Nucleotide substitution causing Pfkelch13 A724E 6
Novelty of Pfkelch13 A724E 6
**Supplementary Figures** 7
Figure S1. Structural characterisation and molecular dynamics simulations of the PfKelch13 A724E mutation. 7
Figure S2. Observed and modelled prevalence of Pfkelch13 in Kaoma, Zambia. 9
Figure S3. Pre- and post-treatment parasite density for day-3 positive patients. 10
**Supplementary Tables** 11
Table S1. Primer sequences and pooling for microsatellite genotyping. 11
Table S2. Number of isolates with Pfkelch13 sequencing results for the two cross-sectional studies conducted in Zambia in 2024. 12
Table S3. Prevalence of Pfkelch13 mutations above 0.5% prevalence across Zambia in 2024. 13
Table S4. Prevalence of Pfkelch13 mutations observed in 2024 across Zambia. 13
Table S5. Prevalence of Pfkelch13 A724E in 2024 in Zambia by province. 14
Table S6. Microsatellite diversity compared between Pfkelch13 A724E and wildtype parasites by locus. 15
Table S7. Mean microsatellite diversity across haplotypes compared between Pfkelch13 A724E and wildtype parasites. 15
Table S8. Prevalence of Pfkelch13 A724E in Kaoma, Western Province. 16
Table S9. Univariable analysis comparing patients retained until day 3 and those lost to follow-up. 16
No significant differences were observed between measured baseline patient characteristics and prevalence of A724E in the two groups. 16
Table S10. Patient characteristics and Pfkelch13 genotyping results for N=98 patients who returned on day 3 post-treatment. 16
Table S11. Univariable analysis of associations between covariates and day-3 positivity. 18
Table S12. Univariable analysis of associations between covariates and day-3 positivity, excluding 16 patients who carried Pfkelch13 mutations other than A724E. 19
Table S13. Multivariable analysis of associations between covariates and day-3 positivity, excluding 16 patients who carried Pfkelch13 mutations other than A724E. 19

Table S14. Within-patient selection of Pfkelch13 A724E. 20
Table S15. Ex vivo growth inhibition for dihydroartemisinin and lumefantrine by Pfkelch13 genotype. 20
**References 21**

#### Author Contributions

M.M., W.L., J.C., C.F., D.J.B., M.H. and J.A.H. designed the study. M.M., B.M., C.C., and K.M. performed the nanopore sequencing. K.M. performed the microsatellite genotyping. M.M., R.O., and C.C. performed the *ex vivo* assays. M.M., M.P., A.S., V.C., T.N., M.Musanje., M.I., C.Chirwa, J.C., I.M., E.S., S.B., J.M., R.C., C.F. and M.H. supervised and collected data in the clinical study. R.Coppée and M-A.S. performed the *in silico* structural analysis. D.M., M.V., C.D., R.P., R.V., F.P.M. and S.P. provided resources and supervision. B.B. and J.A.H. performed the data analysis. J.A.H. wrote the first draft of the manuscript. All authors reviewed and approved the manuscript. M.M, C.F., D.J.B., and J.A.H. decided to publish.

#### Supplementary Methods

##### Nanopore sequencing

We performed targeted nanopore sequencing using the NOMADS-MVP protocol[1]. Briefly, following DNA extraction, an amplicon covering *PfKelch13* from codon 383 to the C-terminus (codon 727) was amplified as part of a 25µL multiplex PCR using KAPA HiFI ReadyMix (Roche, KK2602). PCR products were cleaned at a 0.5X volumetric ratio with AMPure XP beads (Beckman Coulter, A63881) and DNA libraries were prepared for Oxford Nanopore Technologies (ONT) sequencing using the Rapid Barcoding Kit (SQK-RBK114.96) according to manufacturers instructions with the following modifications: 1µL of barcode (RB01-96) was used per sample; 1µL of RA and 2.3µL ADB were combined for adapter ligation; 800-1,200ng of DNA was loaded per R10.4.1 flow cell. Sequencing was performed on a MinION Mk1B device using MinKNOW (v24) and the super-accurate (SUP) basecalling model. A step-by-step protocol can be found on protocols.io.

For nanopore sequencing, FASTQ files were processed using the *Nomadic* pipeline (v0.7.0) which, in brief, maps reads to the *P. falciparum* 3D7 reference genome (PlasmoDB release 67) using Minimap2 (v2.28), calls variants using Delve (v0.2.0), and annotates amino acid changes using Bcftools (v1.20). For quality control, we excluded: (1) isolates with less than 100x mean coverage over the *Pfkelch13* amplicon; (2) isolates with less than 10x the coverage of the negative control; (3) experiments with >100-fold coverage in the negative control; (4) isolates identified as probable duplicates based on a clustering analysis of the *ama1* within-sample allele frequencies.

##### Microsatellite genotyping

Primer sequences, concentrations, and annealing temperatures are described in Table S1. PCR mixtures contained 7.5uL of KAPA HiFi ReadyMix (Roche #KK2602), 2uL of sample DNA, and the indicated microsatellite primer concentration (STable 1) in 15uL total volume. Cycling conditions were: initial denaturation at 95°C for 2 min; followed by forty cycles of denaturation at 95°C for 30 s, annealing at 48°C or 55°C for 30 s, and extension at 60°C for 30 s; with a final extension at 60°C for 1 min. Plates were prepared with amplification products from one or two microsatellite markers and included positive and negative controls to assess consistency and contamination. Capillary electrophoresis was performed by Eurofins Genomics (Ebersberg, Germany) on an ABI 3130 XL instrument with a ROX-500 size standard.

Peaks were called using GeneMapper Software 5 (Applied Biosystems) and isolates with all seven microsatellite markers successfully genotyped (122 of 136, 89.7%) were retained for downstream analysis. Bins were created for each microsatellite marker based on the repeat unit size, and alleles were called for the primary peak using Python.

#### Molecular dynamics simulations

Molecular dynamics simulations were performed on the monomeric state of the wildtype and A724E mutant Kelch protein K13 from AlphaFold (Q8IDQ2), including residues 350 to 726 which contain the BTB and propeller domains. We used the GROMACS[2] package 2025 with the Amber99ss-ILDN force field, which includes improved side-chain torsion potentials for amino acid interactions. The protein systems were immersed in a dodecahedral box containing TIP4P water molecules, with at least 13 Å of separation between the solute and the box's edge. We employed the Particle Mesh Ewald (PME) approach with van der Waals and Coulomb non-bonded interactions truncated at 10 Å. We constrained bond lengths using the LINCS algorithm, which allowed a 2-femtosecond time step in all simulations. The ionization state of the residues was set to be consistent with a neutral pH. For that, Na+ counterions were added by randomly replacing water molecules to ensure the system's overall charge neutrality. The entire system contained approximately 81,000 atoms. To release conflicting contacts, the solvated systems underwent energy minimization using the steepest descent algorithm over 5,000 steps, achieving a maximum force of less than 1,000 kJ/mol/nm. Before molecular dynamics production, each solvated system underwent two-step equilibration. First, the systems were equilibrated for 100 ps in the NVT ensemble at 300 K with V-rescale temperature coupling. Then, the equilibrated systems from the NVT ensemble were treated in the NPT ensemble for 100 ps using the Parrinello-Rahman barostat under an isothermal-isobaric pressure of 1.0 bar. Position restraints were applied to all atoms during the equilibration steps to prevent changes in configuration. Molecular dynamics simulations were run for 200 ns without any restraints. During productions, the V-rescale thermostat coupled with the Parrinello-Rahman barostat were used to maintain the temperature and pressure at 300 K and 1.0 bar, respectively. Frames of the trajectories were stored every 10 ps.

##### Archival sample sequencing

In order to assess the longitudinal prevalence of *Pfkelch13* A724E, we sequenced archival isolates collected as part of three previous studies which included collections in the district of Kaoma, Western Province, Zambia (Figure 3, Figure S2, Table S8). These studies were the Zambia Malaria Indicator Survey in 2018, as well as therapeutic efficacy studies conducted in 2021 and 2023. For each of these studies, all available isolates from Kaoma were sequenced. The studies were approved by the University of Zambia Biomedical Research Ethics Committee (REF No.11-02-18) and ERES Converge (REF No.2021-April-23, REF No.2023-Nov-010). Data from 2024 and 2026 derived from the studies presented in this paper.

### Supplementary Information

#### Nucleotide substitution causing *Pfkelch13* A724E

*Pfkelch13* A724E is caused by a guanine (G) to thymine (T) substitution on the forward strand of chromosome 13 at position 1,724,827 of the *Plasmodium falciparum* 3D7 reference genome, downloaded from PlasmoDB (version 67 from https://plasmodb.org/). This corresponds to a GCA (A) to GAA (E) substitution within codon 724 on the reverse strand, which encodes *Pfkelch13.*

#### Novelty of *Pfkelch13* A724E

We searched public databases for previous reporting of *Pfkelch13* A724E. No changes at codon 724 of *Pfkelch13* were found in the global WorldWide Antimalarial Resistance Network database[3] (accessed 24 March 2026) or the global WHO Malaria Threat Map database[4] (accessed 6 April 2026). In the global MalariaGEN Pf8 data release[5], which includes 33,325 *P. falciparum* whole genome samples, we observed four samples with changes at codon 724, but none were A724E. *Pfkelch13* A724E was not observed in 282 *P. falciparum* whole genome samples collected during the 2018 malaria indicator survey in Zambia[6]. The largest recent study conducted Illumina amplicon sequencing on 2,486 samples collected from across Zambia in 2023[7], but the assay did not cover codon 724.

### Supplementary Figures

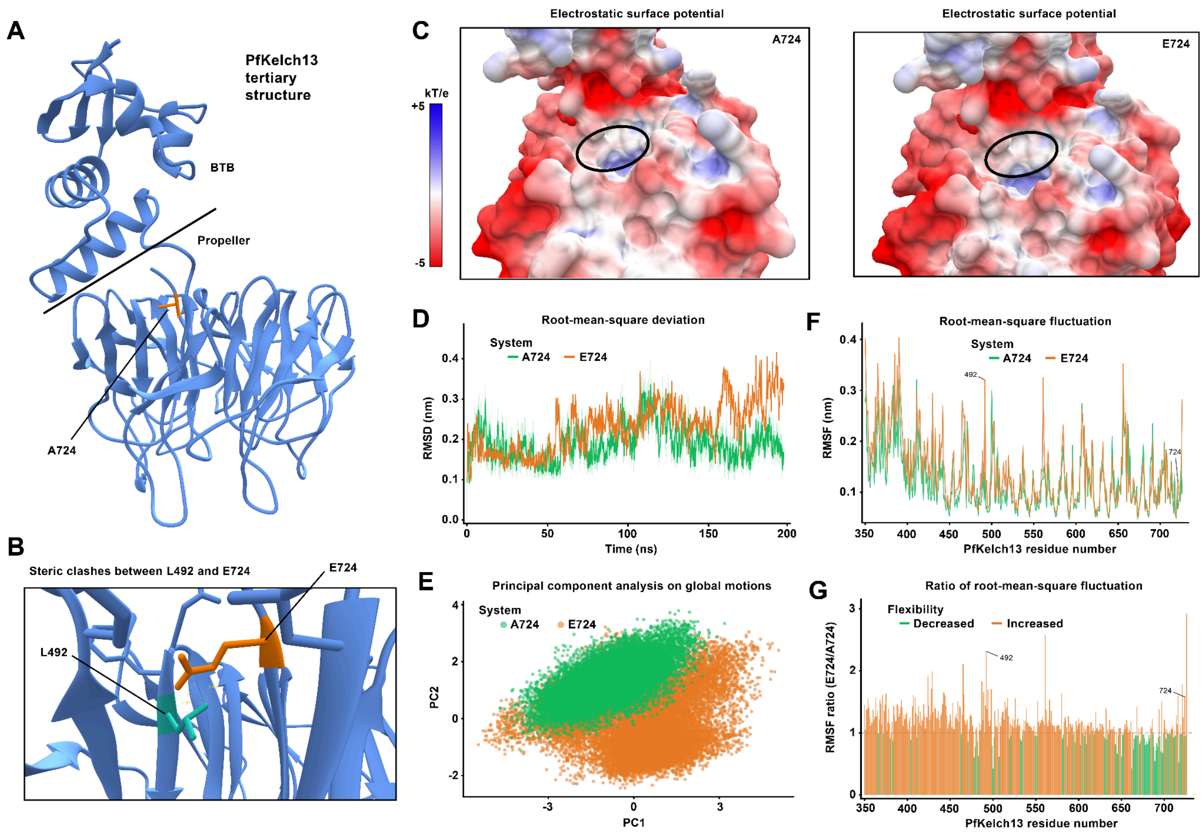


Figure S1. **Structural characterisation and molecular dynamics simulations of the *PfKelch13* A724E mutation.**

**A. Structural architecture and mutation localization.** Three-dimensional representation of the *PfKelch13* protein, highlighting the tertiary organization of the BTB and the C-terminal Kelch-propeller domains. The wild-type residue at position 724 is highlighted in orange; its localization is within the hydrophobic interior of the Kelch-propeller domain. **B. Local steric consequences of the A724E substitution.** The substitution of small, non-polar alanine with bulky, negatively charged glutamic acid induces severe steric hindrance, triggering direct atomic clashes with the neighbouring L492 residue. Beyond physical crowding, the insertion of a hydrophilic, charged carboxylate group into the tightly packed hydrophobic core imposes a severe thermodynamic desolvation penalty, as the glutamate side chain preferentially favours solvent exposure or stabilizing salt-bridge networks rather than a buried hydrophobic environment. **C. Solvent-accessible electrostatic surface potential of both wild-type and mutant structures.** The site of mutation (delineated by the black circle) shows no significant alterations in the global surface charge distribution. This lack of surface electrostatic modification confirms that the E724 residue remains fully sequestered and buried deep within the internal core of the propeller domain, preventing direct interaction with the external solvent. **D. Backbone root-mean-square deviation (RMSD) of wild-type and mutant structures over time.** The wild-type system (green) achieves rapid equilibration and maintains an overall stable conformation throughout the trajectory, fluctuating tightly around a mean value of 0.19 nm. The A724E mutant (orange) exhibits pronounced and progressive structural divergence, demonstrating an upward drift after 150 ns that culminates in peaks exceeding 0.35 nm. The elevated average RMSD of the mutant (0.23 nm vs. 0.19 nm for wild-type) provides evidence of a time-dependent departure from the native fold, signifying global structural destabilization. **E. Principal Component Analysis (PCA) of essential dynamics.** Only the conformational sampling trajectories along the first two principal components (PC1 vs. PC2) are shown. The wild-type protein (green) occupies a highly localized, compact cluster, indicative of a rigid structure confined to a narrow, well-defined native energy well. Conversely, the A724E mutant (orange) displays an expanded and scattered point distribution, reflecting significantly broader conformational freedom. This wider sampling suggests that the mutation compromises structural constraints, forcing the protein to wander across a disorganized landscape of multiple distinct conformational sub-states. **F. Root-mean-square fluctuation (RMSF) profiles mapping local backbone flexibility.** The A724E mutant displays distinct hyper-flexibility peaks compared to the wild-type. A major spike in fluctuations is observed directly around residue L492, capturing the dynamic repercussions of the local steric clashes and electrostatic mismatches introduced by the 724E variant. This local perturbation acts as a destabilizing focal point, propagating structural relaxation through adjacent loop regions and secondary structure elements across the domain. **G. Side-chain root-mean-square fluctuation (RMSF) ratio distribution.** The mutant exhibits an elevated ratio across nearly the entirety of the protein, indicating generalized side-chain hyper-flexibility. This widespread increase in side-chain mobility corroborates the loss of internal packing constraints and suggests a systemic thermodynamic destabilization of the PfKelch13 tertiary structure, a phenotype strongly correlated with accelerated protein degradation and subsequent artemisinin resistance.

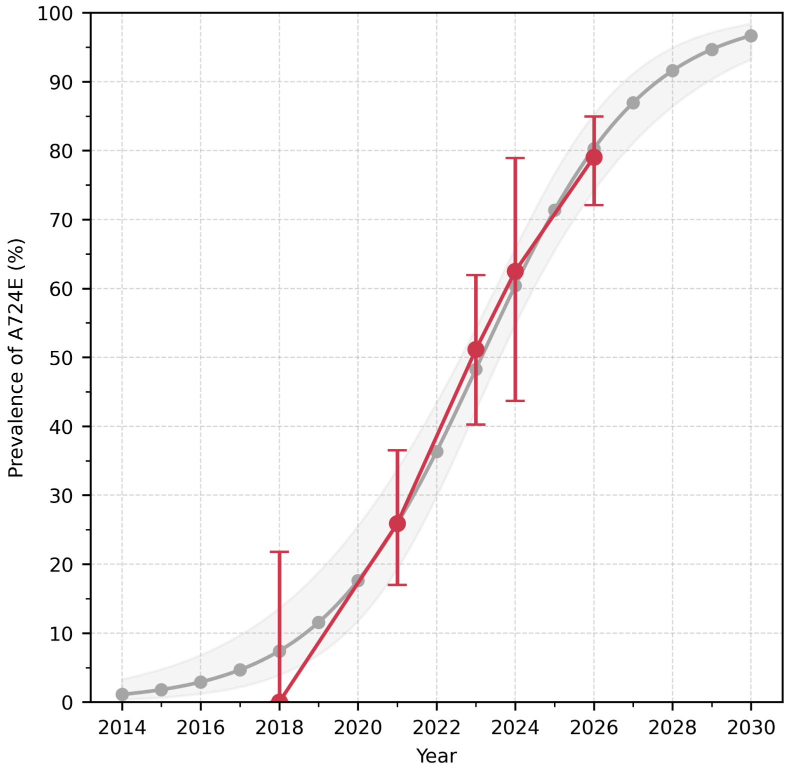


Figure S2**. Observed and modelled prevalence of *Pfkelch13* in Kaoma, Zambia.**

Observed prevalence data are shown in red as in Figure 3. We fit a generalised linear model with a logit link function and binomial outcome variable to the observed prevalence data; point estimates of the A724E selection coefficient and time of 50% prevalence exceedance were obtained with maximum-likelihood estimation. The predicted mean prevalence of A724E is in grey, and the shaded area shows 95% confidence interval.

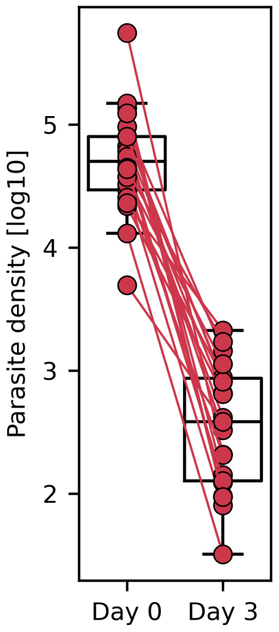


Figure S3**. Pre- and post-treatment parasite density for day-3 positive patients.**

Pre-treatment parasite density (day 0) and post-treatment parasite density (day 3) are shown on a logarithmic scale for *N=21* day-3 positive patients. In these 21 patients, the median parasite density was 50,000 (interquartile range [IQR] 29,256 to 79,355) parasites per cubic millimetre pre-treatment versus 382 (IQR 127 to 861) on day 3. The median decrease in parasite density pre- versus post-treatment was 142-fold (range 10.8 to 2667-fold).

### Supplementary Tables

Table S1. Primer sequences and pooling for microsatellite genotyping.

| Locus (kbp) | Primer | Sequence | Pool | Product (bp) | Repeat Unit |
|---|---|---|---|---|---|
| -0.15 | F | GATTCCCTATCATACGTCATAG | 1 | 224 | AT |
| | R | /56FAM/ACAAGGCGTAAATATTCGTG | | | |
| -3.7 | F | /56FAM/ATGAAAAGGAACATACACAC | 3 | 163 | AT |
| | R | GCACAAATCACCATATTATT | | | |
| -6.36 | F | AATTAGACAGGACTTGTTAATG | 4 | 285 | ATT |
| | R | /56FAM/GGATATGTTATCCAAAATGCA | | | |
| -31.9 | F | /56FAM/AATCAATCATGGAAATTATG | 2 | 208 | AT |
| | R | TTGTTCATTTAAGGGTATTT | | | |
| 15.1 | F | CTTACTTCTAGGAGAATCCT | 1 | 149 | ATT |
| | R | /5HEX/GATAACTACTCATCTCCACA | | | |
| 31 | F | TATAAGATTCGTTATTACATG | 3 | 294 | GTTCATTTT |
| | R | /5HEX/TATAAGATTCGTTATTACATG | | | |
| 72.3 | F | ATGGCTCTATAATAGGAAGG | 2 | 239 | TA |
| | R | /5HEX/TGCAAACTTTTTAGACTTTT | | | |

Pairs of forward and reverse primers with different fluorophores were pooled. For all reactions we used 0.7uL of primer and 55ºC annealing; except for the 31kbp locus, where we used 1.1uL and 48ºC.

Table S2. Number of isolates with *Pfkelch13* sequencing results for the two cross-sectional studies conducted in Zambia in 2024.

| Study | Province | Failing – n (%) | Passing – n (%) | Overall – n |
|---|---|---|---|---|
| HRP23 | Central | 33 (23) | 110 (77) | 143 |
| HRP23 | Copperbelt | 35 (13) | 239 (87) | 274 |
| HRP23 | Eastern | 45 (11) | 347 (89) | 392 |
| HRP23 | Luapula | 93 (20) | 377 (80) | 470 |
| HRP23 | Lusaka | 5 (6) | 82 (94) | 87 |
| HRP23 | Muchinga | 10 (7) | 138 (93) | 148 |
| HRP23 | Northern | 2 (4) | 49 (96) | 51 |
| HRP23 | North-Western | 34 (19) | 146 (81) | 180 |
| HRP23 | Western | 40 (18) | 179 (82) | 219 |
| MIS2024 | Central | 11 (37) | 19 (63) | 30 |
| MIS2024 | Copperbelt | 10 (21) | 37 (79) | 47 |
| MIS2024 | Eastern | 25 (42) | 34 (58) | 59 |
| MIS2024 | Luapula | 57 (66) | 29 (34) | 86 |
| MIS2024 | Lusaka | 7 (47) | 8 (53) | 15 |
| MIS2024 | Muchinga | 7 (18) | 31 (82) | 38 |
| MIS2024 | Northern | 16 (25) | 48 (75) | 64 |
| MIS2024 | North-Western | 13 (25) | 38 (75) | 51 |
| MIS2024 | Southern | 0 (0) | 1 (100) | 1 |
| MIS2024 | Western | 6 (14) | 38 (86) | 44 |
| HRP23 | All | 297 (15) | 1667 (85) | 1964 |
| MIS2024 | All | 152 (35) | 283 (65) | 435 |
| All | All | 449 (19) | 1950 (81) | 2399 |

HRP23 refers to the molecular study; MIS2024 the malaria indicator survey.

Table S3. Prevalence of *Pfkelch13* mutations above 0.5% prevalence across Zambia in 2024.

| Mutation | Prevalence (95% CI) – % | N | Mutant – n | Mixed – n | Mutant or Mixed – n |
|---|---|---|---|---|---|
| P441L[b] | 2.6 (1.9-3.4) | 1948 | 28 | 22 | 50 |
| C473F | 0.6 (0.3-1.0) | 1950 | 4 | 7 | 11 |
| G533A | 0.8 (0.4-1.3) | 1950 | 7 | 8 | 15 |
| P574L[a] | 0.5 (0.2-0.9) | 1950 | 2 | 7 | 9 |
| A578S | 1.0 (0.6-1.5) | 1950 | 6 | 13 | 19 |
| R622T | 0.8 (0.5-1.3) | 1950 | 8 | 8 | 16 |
| P667A | 1.5 (1.0-2.2) | 1949 | 10 | 20 | 30 |
| A724E | 11.7 (10.4-13.3) | 1949 | 111 | 118 | 229 |
| Other[c] | 4.3 (4.0-4.7) | 1950 | 19 | 65 | 84 |
| Total | 23.7 (23.1-24.4) | 1950 | 195 | 268 | 463 |

*Pfkelch13* was successfully sequenced for a total of *N=1950* isolates. Only non-synonymous mutations with frequency ≥0.5% are shown. Note that *N* is slightly less than 1950 for three mutations (P441L, P667A, A724E); this is caused by cases where isolates have passed overall quality control, but failed mutation-specific quality filters (read strand bias and base Phred scores).

[a]WHO validated ART-R marker.

[b]WHO candidate ART-R marker.

[c]Includes 34 other *Pfkelch13* mutations with frequencies <0.5%; see Table S2.

Table S4. Prevalence of *Pfkelch13* mutations observed in 2024 across Zambia.

| Mutation | Prevalence (95% CI) – % | N | Mutant – n | Mixed – n | Mutant or Mixed – n |
|---|---|---|---|---|---|
| D389E | 0.3 (0.1-0.7) | 1934 | 0 | 6 | 6 |
| S400I | 0.1 (0.0-0.3) | 1948 | 0 | 1 | 1 |
| R411K | 0.3 (0.1-0.7) | 1946 | 0 | 6 | 6 |
| R411E | 0.1 (0.0-0.4) | 1946 | 0 | 2 | 2 |
| P419S | 0.1 (0.0-0.3) | 1949 | 1 | 0 | 1 |
| S425C | 0.1 (0.0-0.3) | 1949 | 0 | 1 | 1 |
| L428F | 0.1 (0.0-0.4) | 1950 | 0 | 2 | 2 |
| P441L[b] | 2.6 (1.9-3.4) | 1948 | 28 | 22 | 50 |
| P441R | 0.1 (0.0-0.4) | 1948 | 0 | 2 | 2 |
| P443S | 0.1 (0.0-0.4) | 1950 | 2 | 0 | 2 |
| C473F | 0.6 (0.3-1.0) | 1950 | 4 | 7 | 11 |
| G484V | 0.1 (0.0-0.4) | 1949 | 0 | 2 | 2 |
| A504V | 0.1 (0.0-0.3) | 1949 | 0 | 1 | 1 |
| R513S | 0.2 (0.0-0.4) | 1950 | 0 | 3 | 3 |
| C532S | 0.1 (0.0-0.4) | 1950 | 0 | 2 | 2 |
| G533A | 0.8 (0.4-1.3) | 1950 | 7 | 8 | 15 |
| R539I | 0.3 (0.1-0.7) | 1949 | 2 | 4 | 6 |
| C542F | 0.1 (0.0-0.3) | 1950 | 0 | 1 | 1 |
| S550Y | 0.1 (0.0-0.4) | 1949 | 0 | 2 | 2 |
| D559Y | 0.1 (0.0-0.4) | 1950 | 0 | 2 | 2 |
| M562I | 0.1 (0.0-0.3) | 1950 | 1 | 0 | 1 |
| A569V | 0.1 (0.0-0.4) | 1949 | 0 | 2 | 2 |

| | | | | | |
|---|---|---|---|---|---|
| T573A | 0.1 (0.0-0.4) | 1950 | 1 | 1 | 2 |
| T573I | 0.1 (0.0-0.4) | 1950 | 0 | 2 | 2 |
| P574L[a] | 0.5 (0.2-0.9) | 1950 | 2 | 7 | 9 |
| A578S | 1.0 (0.6-1.5) | 1950 | 6 | 13 | 19 |
| V589I | 0.1 (0.0-0.3) | 1949 | 1 | 0 | 1 |
| R597I | 0.1 (0.0-0.4) | 1950 | 0 | 2 | 2 |
| Q613E | 0.2 (0.0-0.4) | 1950 | 0 | 3 | 3 |
| R622T | 0.8 (0.5-1.3) | 1950 | 8 | 8 | 16 |
| A626E | 0.2 (0.0-0.4) | 1950 | 0 | 3 | 3 |
| V637I | 0.1 (0.0-0.4) | 1950 | 1 | 1 | 2 |
| V650F | 0.1 (0.0-0.4) | 1950 | 0 | 2 | 2 |
| G665V | 0.1 (0.0-0.3) | 1947 | 0 | 1 | 1 |
| V666L | 0.2 (0.0-0.4) | 1950 | 2 | 1 | 3 |
| P667S | 0.4 (0.2-0.8) | 1949 | 5 | 3 | 8 |
| P667A | 1.5 (1.0-2.2) | 1949 | 10 | 20 | 30 |
| A675V[a] | 0.3 (0.1-0.6) | 1950 | 2 | 3 | 5 |
| W706L | 0.1 (0.0-0.4) | 1950 | 0 | 2 | 2 |
| S711Y | 0.1 (0.0-0.4) | 1950 | 1 | 1 | 2 |
| S711P | 0.1 (0.0-0.3) | 1950 | 0 | 1 | 1 |
| A724E | 11.7 (10.4-13.3) | 1949 | 111 | 118 | 229 |

[a]WHO validated artemisinin partial resistance marker
[b]WHO candidate artemisinin partial resistance marker

Table S5. Prevalence of *Pfkelch13* A724E in 2024 in Zambia by province.

| **Province** | **Area – km$^2$ (000)** | **Prevalence (95% CI) – %** | **N** | **Mutant – n** | **Mixed – n** | **Mutant or Mixed – n** |
|---|---|---|---|---|---|---|
| Western | 127 | 52.1 (45.2-58.9) | 217 | 54 | 59 | 113 |
| North-Western | 125 | 51.1 (43.6-58.5) | 184 | 46 | 48 | 94 |
| Southern | 84 | 0.0 (0.0-97.5) | 1 | 0 | 0 | 0 |
| Copperbelt | 31 | 3.3 (1.5-6.1) | 275 | 3 | 6 | 9 |
| Luapula | 50 | 0.0 (0.0-0.9) | 406 | 0 | 0 | 0 |
| Lusaka | 23 | 8.9 (3.9-16.8) | 90 | 6 | 2 | 8 |
| Central | 95 | 3.1 (0.9-7.7) | 129 | 1 | 3 | 4 |
| Northern | 78 | 0.0 (0.0-3.7) | 97 | 0 | 0 | 0 |
| Muchinga | 69 | 0.6 (0.0-3.3) | 169 | 1 | 0 | 1 |
| Eastern | 69 | 0.0 (0.0-1.0) | 381 | 0 | 0 | 0 |

Table S6. Microsatellite diversity compared between *Pfkelch13* A724E and wildtype parasites by locus.

| Province | Locus — kbp | 724A — N | 724A — $H_e$ | 724E — N | 724E — $H_e$ | $\Delta H_e$ | P Value |
|---|---|---|---|---|---|---|---|
| North-Western | -31.9 | 31 | 0.91 | 22 | 0.26 | 0.65 | <.001 |
| North-Western | -6.36 | 31 | 0.77 | 22 | 0.17 | 0.59 | <.001 |
| North-Western | -3.7 | 31 | 0.89 | 22 | 0 | 0.89 | <.001 |
| North-Western | -0.15 | 31 | 0.79 | 22 | 0.25 | 0.54 | <.001 |
| North-Western | 15 | 31 | 0.54 | 22 | 0.09 | 0.44 | .0089 |
| North-Western | 31 | 31 | 0.61 | 22 | 0.09 | 0.52 | <.001 |
| North-Western | 72.3 | 31 | 0.93 | 22 | 0.81 | 0.12 | .0018 |
| Western | -31.9 | 42 | 0.91 | 25 | 0.42 | 0.49 | <.001 |
| Western | -6.36 | 42 | 0.69 | 25 | 0.35 | 0.35 | <.001 |
| Western | -3.7 | 42 | 0.74 | 25 | 0.08 | 0.66 | <.001 |
| Western | -0.15 | 42 | 0.86 | 25 | 0.08 | 0.78 | <.001 |
| Western | 15 | 42 | 0.76 | 25 | 0 | 0.76 | <.001 |
| Western | 31 | 42 | 0.61 | 25 | 0.08 | 0.53 | <.001 |
| Western | 72.3 | 42 | 0.87 | 25 | 0.69 | 0.18 | .0043 |
| Both | -31.9 | 73 | 0.91 | 47 | 0.34 | 0.57 | <.001 |
| Both | -6.36 | 73 | 0.73 | 47 | 0.26 | 0.46 | <.001 |
| Both | -3.7 | 73 | 0.82 | 47 | 0.04 | 0.77 | <.001 |
| Both | -0.15 | 73 | 0.85 | 47 | 0.16 | 0.69 | <.001 |
| Both | 15 | 73 | 0.7 | 47 | 0.04 | 0.66 | <.001 |
| Both | 31 | 73 | 0.61 | 47 | 0.08 | 0.52 | <.001 |
| Both | 72.3 | 73 | 0.9 | 47 | 0.74 | 0.16 | <.001 |

724E is mutant and 724A is wildtype. $H_e$ is the expected heterozygosity. P values were calculated using a permutation test with 1,000 iterations.

Table S7. Mean microsatellite diversity across haplotypes compared between *Pfkelch13* A724E and wildtype parasites.

| Province | 724A — N | 724E — N | 724A — $H_e$ | 724AE— $H_e$ | $\Delta H_e$ | P Value |
|---|---|---|---|---|---|---|
| Both | 73 | 47 | 0.79 | 0.24 | 0.55 | <.001 |
| North-Western | 31 | 22 | 0.78 | 0.24 | 0.54 | <.001 |
| Western | 42 | 25 | 0.78 | 0.24 | 0.54 | <.001 |

724E is mutant and 724A is wildtype. $H_e$ is the expected heterozygosity. P values were calculated using a permutation test with 1,000 iterations.

Table S8. Prevalence of *Pfkelch13* A724E in Kaoma, Western Province.

| Year | Prevalence (95% CI) – % | N | Mixed – n | Mutant – n | Mutant or Mixed – n |
|---|---|---|---|---|---|
| 2018 | 0.0 (0.0-21.8) | 15 | 0 | 0 | 0 |
| 2021 | 25.9 (17.0-36.5) | 85 | 11 | 11 | 22 |
| 2023 | 51.1 (40.2-61.9) | 88 | 26 | 19 | 45 |
| 2024 | 62.5 (43.7-78.9) | 32 | 14 | 6 | 20 |
| 2026 | 79.0 (72.1-84.9) | 167 | 79 | 53 | 132 |

Table S9. Univariable analysis comparing patients retained until day 3 and those lost to follow-up.

| | Retained | Lost | Test | P |
|---|---|---|---|---|
| N | 103 | 85 | – | – |
| Median Age (IQR) – yr | 12.0 (6.2 – 17.0) | 11.5 (6.0 – 20.0) | Mann-Whitney U | 0.853 |
| Median parasite density (IQR) – $mm^3$ | 26.4 (14.0 – 54.8) | 29.3 (9.7 – 52.9) | Mann-Whitney U | 0.564 |
| Female sex – n (%) | 54 (52.4) | 39 (45.9) | $\chi^2$ | 0.503 |
| Fever – n (%) | 23 (22.3) | 14 (16.5) | $\chi^2$ | 0.362 |
| Prevalence of A724E (95% CI) – n (%) | 76.5 (66.9 – 84.5) | 81.8 (71.4 – 89.7) | $\chi^2$ | 0.507 |

No significant differences were observed between measured baseline patient characteristics and prevalence of A724E in the two groups.

Table S10. Patient characteristics and *Pfkelch13* genotyping results for *N=98* patients who returned on day 3 post-treatment.

| Patient Characteristics | | Pre-treatment (Day 0) | | Post-treatment (Day 3) | |
|---|---|---|---|---|---|
| Sex | Fever | *Pfkelch13* Genotype | *Pfkelch13* Total WSAF | *Pfkelch13* Total WSAF | Parasite Positive |
| F | | WT | 0 | | FALSE |
| F | No | A724E | 1 | 1 | TRUE |
| F | No | A724E | 0.28 | | FALSE |
| M | Yes | A724E | 0.8 | Failed | TRUE |
| M | No | A724E | 0.18 | | FALSE |
| F | No | A724E | 1 | | FALSE |
| F | No | A724E | 1 | 1 | TRUE |
| M | No | WT | 0 | | FALSE |
| F | Yes | WT | 0 | | FALSE |
| M | Yes | WT | 0 | | FALSE |
| F | No | A724E | 1 | | FALSE |
| M | No | P667S+A724E | 0.8 | | FALSE |
| M | No | WT | 0 | | FALSE |
| F | Yes | A724E | 0.98 | 1 | TRUE |
| F | Yes | P667A+A724E | 1 | | FALSE |
| F | Yes | WT | 0 | | FALSE |

| | | | | | |
|---|---|---|---|---|---|
| M | Yes | A724E | 0.25 | 0.95 | TRUE |
| F | No | A724E | 0.81 | | FALSE |
| M | Yes | G533A | 1 | | FALSE |
| M | No | A724E | 1 | | FALSE |
| M | No | A724E | 0.1 | | FALSE |
| M | No | A724E | 0.74 | 1 | TRUE |
| F | Yes | A724E | 1 | 1 | TRUE |
| M | No | A724E | 1 | | FALSE |
| M | No | A724E | 1 | | FALSE |
| F | No | A724E | 0.49 | 1 | TRUE |
| M | No | A724E | 1 | | FALSE |
| F | | WT | 0 | | FALSE |
| M | | P667A+A724E | 0.95 | | FALSE |
| M | | WT | 0 | | FALSE |
| F | No | A724E | 1 | | FALSE |
| M | No | P667A | 1 | | FALSE |
| F | No | WT | 0 | | FALSE |
| M | No | WT | 0 | | FALSE |
| M | No | A724E | 0.08 | | FALSE |
| F | No | A724E | 1 | 0.99 | TRUE |
| F | No | WT | 0 | | FALSE |
| M | No | P667A+A724E | 0.98 | | FALSE |
| F | No | WT | 0 | | FALSE |
| F | No | A724E | 0.19 | | FALSE |
| F | No | P667A+A724E | 0.99 | 0.99 | TRUE |
| M | No | A724E | 1 | Failed | TRUE |
| M | No | A724E | 1 | | FALSE |
| F | No | A724E | 1 | | FALSE |
| M | No | P667A+A724E | 0.84 | | FALSE |
| M | No | WT | 0 | | FALSE |
| F | No | A724E | 1 | | FALSE |
| F | No | A724E | 0.94 | | FALSE |
| F | Yes | A724E | 0.33 | | FALSE |
| M | No | A724E | 0.93 | | FALSE |
| F | No | P441L+A724E | 0.68 | | FALSE |
| M | No | A724E | 0.95 | | FALSE |
| M | No | R622T+A724E | 0.73 | | FALSE |
| M | No | A724E | 0.67 | | FALSE |
| M | No | A724E | 1 | | FALSE |
| M | No | A724E | 0.68 | | FALSE |
| F | No | WT | 0 | | FALSE |
| F | No | A675V+A724E | 0.67 | 1 | TRUE |
| F | Yes | A675V+A724E | 0.69 | | FALSE |
| F | No | WT | 0 | | FALSE |
| F | No | A724E | 1 | | FALSE |
| F | No | R622T+A724E | 0.2 | Failed | TRUE |
| F | No | A724E | 1 | | FALSE |
| F | Yes | A724E | 0.99 | | FALSE |
| M | Yes | A724E | 1 | 1 | TRUE |
| M | Yes | A724E | 1 | | FALSE |
| M | No | A724E | 1 | | FALSE |
| F | No | A724E | 1 | 1 | TRUE |
| M | No | A724E | 0.05 | | FALSE |
| M | No | A724E | 0.23 | | FALSE |
| F | No | A724E | 1 | | FALSE |
| F | Yes | A724E | 0.42 | 0.96 | TRUE |
| M | No | A724E | 1 | | FALSE |
| F | No | A724E | 1 | | FALSE |
| M | No | WT | 0 | | FALSE |
| F | No | A724E | 0.41 | | FALSE |
| M | No | A724E | 1 | 0.99 | TRUE |

| | | | | | |
|---|---|---|---|---|---|
| F | No | A724E | 1 | 1 | TRUE |
| F | No | A724E | 0.98 | | FALSE |
| F | No | A724E | 1 | | FALSE |
| M | No | A724E | 0.99 | Failed | TRUE |
| M | Yes | A724E | 1 | | FALSE |
| F | Yes | A724E | 0.96 | | FALSE |
| F | No | WT | 0 | | FALSE |
| M | No | A724E | 1 | | FALSE |
| F | No | A724E | 0.88 | | FALSE |
| M | Yes | A724E | 0.48 | | FALSE |
| F | No | A724E | 1 | | FALSE |
| F | No | A724E | 1 | | FALSE |
| F | No | P667A | 1 | | FALSE |
| F | No | A724E | 0.35 | 0.62 | TRUE |
| M | Yes | A724E | 0.99 | | FALSE |
| M | Yes | A724E | 0.8 | | FALSE |
| M | No | WT | 0 | | FALSE |
| M | No | WT | 0 | | FALSE |
| F | Yes | P667S+A724E | 1 | 1 | TRUE |
| M | Yes | P667S | 1 | | FALSE |
| F | | A724E | 1 | | FALSE |

For each patient, the total within-sample alternative allele frequency (WSAF) of all *Pfkelch13* mutations is indicated on day 0, and if the patient was day-3 positive, on day 3. The total WSAF is the proportion of parasites within the patient that have a non-wildtype *Pfkelch13* allele. Four day-3 positive patients failed sequencing post-treatment. A total of 16 patients carried *Pfkelch13* mutations other than A724E. WT is wildtype.

#### Table S11. Univariable analysis of associations between covariates and day-3 positivity.

| | Negative on Day 3 | Positive on Day 3 | Test | P |
|---|---|---|---|---|
| N | 77 | 21 | – | – |
| Median Age (IQR) – yr | 12.0 (7.0 - 18.0) | 12.0 (6.0 - 16.0) | Mann-Whitney U | 0.608 |
| Median parasite density (IQR) – $mm^3$ | 25.0 (12.8 - 48.7) | 50.0 (29.3 - 79.4) | Mann-Whitney U | 0.004 |
| Female sex – n (%) | 37 (48.1) | 14 (66.7) | $X^2$ | 0.205 |
| Fever – n (%) | 15 (19.5) | 7 (33.3) | $X^2$ | 0.371 |
| 724A (Wildtype) | 23 (29.9) | 0 (0.0) | Fisher's Exact | 0.003 |
| 724E (Mutant) | 54 (70.1) | 21 (100.0) | | |

All *N=98* patients with successful pre-treatment genotyping of *Pfkelch13* included. Median parasite density and the genotype status at A724E are significant.

Table S12. Univariable analysis of associations between covariates and day-3 positivity, excluding 16 patients who carried *Pfkelch13* mutations other than A724E.

| | Negative on Day 3 | Positive on Day 3 | Test | P |
|---|---|---|---|---|
| N | 65 | 17 | – | – |
| Median Age (IQR) – yr | 11.5 (7.0 - 18.0) | 11.0 (6.0 - 16.0) | Mann-Whitney U | 0.534 |
| Median parasite density (IQR) – $mm^3$ | 25.0 (15.0 - 48.7) | 50.0 (32.6 - 66.4) | Mann-Whitney U | 0.005 |
| Female sex – n (%) | 33 (50.8) | 10 (58.8) | $X^2$ | 0.749 |
| Fever – n (%) | 11 (16.9) | 6 (35.3) | $X^2$ | 0.233 |
| 724A (Wildtype) | 19 (29.2) | 0 (0.0) | Fisher's Exact | 0.009 |
| 724E (Mutant) | 46 (70.8) | 17 (100.0) | | |

A total of *N=82* patients were included, all of which carried either wild-type *Pfkelch13*, or *Pfkelch13* A724E as a mutant or mixed genotype. The 16 patients with other *Pfkelch13* mutations carried P441L, G533A, P667A, P667S or A675V; see Table S10. Median parasite density and the genotype status at A724E remain significant.

Table S13. Multivariable analysis of associations between covariates and day-3 positivity, excluding 16 patients who carried *Pfkelch13* mutations other than A724E.

| Covariate | Coefficient (SE) | Odds Ratio (95% CI) | P |
|---|---|---|---|
| Age | -0.01 (0.03) | 1.0 (0.9 – 1.0) | 0.678 |
| Sex | -0.5 (0.58) | 0.6 (0.2 – 2.0) | 0.408 |
| Fever | 0.48 (0.65) | 1.6 (0.4 – 6.0) | 0.474 |
| Parasite density [log10] | 2.11 (0.89) | 8.2 (1.5 – 68.2) | 0.013 |
| *Pfkelch13 A724E* | 2.44 (1.41) | 11.5 (1.2 – 1574.7) | 0.028 |

The same analysis as presented in Table 1 was performed, but with 16 patients who carried other *Pfkelch13* mutations removed.

Table S14. Within-patient selection of *Pfkelch13* A724E.

| Fever | Sex | Day 0 – Parasite Density | Day 0 – Parasite Density | Day 0 – Pfkelch13 | Day 0 – Total WSAF | Day 3 – Total WSAF | Day 0 – A724E WSAF | Day 3 – A724E WSAF | Change in A724E WSAF? |
|---|---|---|---|---|---|---|---|---|---|
| Yes | F | 66427 | 861 | A724E | 0.98 | 1.00 | 0.98 | 1.00 | increase |
| Yes | M | 553209 | 207 | A724E | 0.25 | 0.95 | 0.25 | 0.95 | increase |
| No | M | 13000 | 32 | A724E | 0.74 | 1.00 | 0.74 | 1.00 | increase |
| Yes | F | 65000 | 328 | A724E | 1.00 | 1.00 | 1.00 | 1.00 | |
| No | F | 60000 | 414 | A724E | 0.49 | 1.00 | 0.49 | 1.00 | increase |
| No | F | 50000 | 123 | A724E | 1.00 | 0.99 | 1.00 | 0.99 | |
| No | F | 4941 | 382 | P667A +A724E | 0.99 | 0.99 | 0.55 | 0.56 | +/- 2% |
| No | F | 29256 | 80 | A675V +A724E | 0.67 | 1.00 | 0.20 | 0.85 | increase |
| Yes | M | 32618 | 206 | A724E | 1.00 | 1.00 | 1.00 | 1.00 | |
| No | F | 54595 | 653 | A724E | 1.00 | 1.00 | 1.00 | 1.00 | |
| Yes | F | 148515 | 95 | A724E | 0.42 | 0.96 | 0.42 | 0.96 | increase |
| No | M | 44537 | 127 | A724E | 1.00 | 0.99 | 1.00 | 0.99 | |
| No | F | 25405 | 874 | A724E | 1.00 | 1.00 | 1.00 | 1.00 | |
| No | F | 123423 | 94 | A724E | 0.35 | 0.62 | 0.35 | 0.62 | increase |
| Yes | F | 79355 | 813 | P667S +A724E | 1.00 | 1.00 | 0.02 | 0.03 | +/- 2% |
| No | F | 37333 | 1127 | A724E | 1.00 | 1.00 | 1.00 | 1.00 | |
| No | F | 43231 | 1705 | A724E | 1.00 | 1.00 | 1.00 | 1.00 | |

Shown are *N=17* day-3 positive patients with successful post-treatment *Pfkelch13* sequencing. Total WSAF is the total within-sample alternative allele frequency of all *Pfkelch13* mutations; A724E WSAF is just for A724E. Two patients had similar A724E WSAF pre- and post-treatment, but in both cases, another *Pfkelch13* A724E was present within the patient (P667A or P677S).

Table S15. *Ex vivo* growth inhibition for dihydroartemisinin and lumefantrine by *Pfkelch13* genotype.

| | 724A | 724E | Test | P |
|---|---|---|---|---|
| N | 20 | 88 | – | – |
| DHA IC50 – nM (IQR) | 0.5 (0.3 – 1.6) | 0.7 (0.3 – 1.1) | Mann-Whitney U | 0.684 |
| Lumefantrine IC50 – nM (IQR) | 4.9 (4.1 – 8.9) | 6.1 (4.2 – 10.8) | Mann-Whitney U | 0.445 |

IC50 is half-maximal inhibitory concentration estimated by fitting a four-parameter Hill Equation to the dose-response curves of two replicate experiments.
